# Increasing HIV treatment literacy among people living with HIV using a novel health communication aid: Evidence from KwaZulu Natal province, South Africa

**DOI:** 10.1101/2023.06.15.23291430

**Authors:** Caroline Govathson, Neo Ndlovu, Letitia Rambally-Greener, Laura Schmucker, Candice Chetty-Makkan, Jacqui Miot, Harsha Thirumurthy, Sophie Pascoe, Shawn Malone, Alison Buttenheim

**Affiliations:** Health Economics and Epidemiology Research Office, Faculty of Health Sciences, University of the Witwatersrand, Johannesburg, South Africa; Population Services International (PSI); Department of Medical Ethics and Health Policy, Perelman School of Medicine, University of Pennsylvania, Philadelphia, PA, USA; Department of Family and Community Health, School of Nursing, University of Pennsylvania, Philadelphia, PA, USA

## Abstract

**Introduction:** Effective health communication is important for promoting adherence to antiretroviral therapy (ART). During counselling sessions with people living with HIV (PLHIV) who are initiating or re-initiating ART, we assessed whether a simple visual aid using bead bottles to explain the concept of viral suppression resulted in changes in HIV treatment literacy.

**Methods:** At three public sector clinics in KwaZulu-Natal KZN) province, South Africa, we enrolled adults who tested HIV-positive and were newly initiating ART or re-engaging in HIV care. Trained HIV counsellors used bottles with coloured beads (“B-OK bottles”) to explain concepts related to viral load, viral suppression, and undetectable=Untransmittable (U=U). We assessed participants’ knowledge, attitudes, and perceptions about ART before and after counselling.

**Results:** Between November 2022 and January 2023, we enrolled 80 PLHIV. Participants’ median age was 32 years (IQR: 24–41) and 58% were male. After receiving counselling with the B-OK bottles, understanding of U=U increased from 6% to 99% and understanding of ‘viral suppression’ increased from 20% to 99%. Confidence in the protective effects of ART increased (64% to 100% for one’s own health; 58% to 94% for transmission to partners) and was observed among participants both initiating ART and re-engaging in care. The number of participants agreeing that viral suppression means their sexual partners are safe from HIV even without condoms increased from 14% to 93% p-value =0.0. However, 65% still expressed worry that ART does not completely eliminate the risk of sexually transmitting HIV.

**Conclusions:** Use of B-OK bottles during ART counselling was acceptable and increased HIV treatment literacy. While there was no significant effect on level of confidence in complete elimination of transmission risk when virally suppressed, we observed a significant reduction in concern about transmitting HIV and an increase in confidence in the protective effects of ART.

**Clinical Trial Number (SANCTR):** DOH-27-092022-8067

## Introduction

Despite scientific evidence about the therapeutic and prevention benefits of antiretroviral therapy (ART), awareness of these benefits remains low in much of sub-Saharan Africa (SSA)(1,2). The prevention benefits of ART in particular have been substantially underestimated in multiple segments of the population, including people living with HIV (PLHIV) and healthcare providers (3,4). Several studies conducted in South Africa, which has the world’s largest HIV epidemic, show that individuals have limited comprehension of the meaning of viral suppression and substantially underestimate the effects of ART on viral load and transmission risk (5,6) .

While increased treatment literacy has the potential to improve ART uptake and retention in care among PLHIV, few evaluations have been conducted on interventions that disseminate information about ART, including the message of “undetectable=untransmittable” (U=U). The most commonly evaluated interventions, including motivational interviewing, health promotion programs, group or individual recipient of care counselling, and active visualisation, have had varying results (7,8,9).

Visual aids have the potential to convey health information in an accessible way to individuals with low numeracy or literacy; this can be particularly effective for facilitating discussions of complex numerical information, such as the percentage reduction in transmission risk due to ART. Visual aids allow for the use of narrative, metaphor, and analogy in HIV treatment adherence counselling (10,11) and can also help recalibrate flawed or outdated mental models about the effects of ART on HIV viral loads and transmission risk (12,13).

The B-OK bead bottles **(Figure 1)** are a simple visual aid that is designed to assist healthcare workers improve HIV treatment literacy among care recipients. Developed by Population Services International (PSI) in collaboration with Matchboxology in South Africa in 2021, using a human-centred design process, the bottles are a conversation starter for nurses and counsellors, assisting them with communicating the importance of treatment adherence, the benefits of viral suppression, and the “good news” of U=U and treatment as prevention. This study examines changes in treatment literacy among PLHIV in South Africa who were introduced to the B-OK bottles during ART initiation or re-engagement in care.

**Figure 1:**
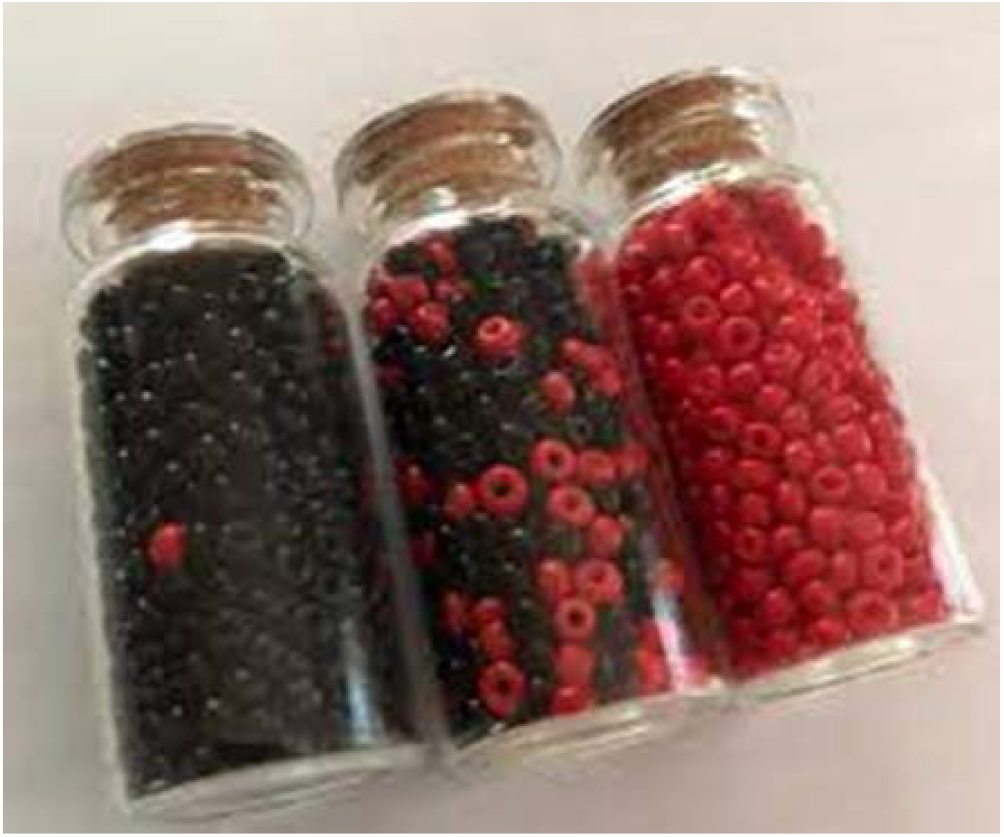
B-OK bead bottles

## Methods

At three public sector clinics in KwaZulu-Natal, South Africa, we enrolled adults who tested HIV-positive and were newly initiating ART or were re-engaging in HIV care after a treatment interruption of at least 90 days. In addition to the standard post-test counselling that all clients receive at the clinics, trained HIV counsellors introduced the three bottles with coloured beads (B-OK bottles, Fig. 1) and used the bottles to explain the meaning of viral load, the effect of ART on viral suppression, and the reduction in transmission risk as a result of viral suppression (i.e., U=U).

Before and immediately after introducing the B-OK bottles, counsellors administered a brief survey that assessed participants’ demographic characteristics and level of treatment literacy. Treatment literacy was measured using nine questions about the need for timely ART initiation, the effect of ART on viral load and the effect of viral suppression on transmission risk (Appendix 1). Participants’ responses to the questions were assigned a score of 1 for correct responses, 0 for “not sure” responses, and -1 for incorrect responses. We then calculated a treatment literacy index score that was the sum of the scores from the 9 items, with a maximum value of 9. The survey also assessed participants’ overall understanding of HIV treatment and their awareness of the concepts of “viral suppression” and “U=U”.

We compared differences in treatment literacy index scores measured before and after exposure to the B-OK bottles using Wilcoxon sign-rank test. This statistical method was chosen due to the paired nature of the data and that the data was non-parametric as determined by a Shapiro-Wilk test. The study was approved by the research ethics committees at the University of the Witwatersrand # M220707 and University of Pennsylvania # 852087.

## Results

Between November 14, 2022 and January 18, 2023, we screened 90 adult PLHIV at study clinics. Ten PLHIV were excluded due to age, ill health or refusal to participate. Among the 80 participants who were enrolled, 23 were newly initiating ART and 55 were re-engaging in care after a treatment interruption of at least 90 days, 2 preferred not to tell us whether they were newly initiating ART or re-engaging with care after a treatment interruption. Median age was 32 years (IQR: 24–41), 46 (58%) were male, 20 (25%) were either married or in a relationship, and 63 (79%) had completed primary school **(Table 1)**.

**Table 1.** Participant characteristics.

| Variable | Categories | All participants | Newly initiating ART | Unknown ART status** | Re-engagement* |
| --- | --- | --- | --- | --- | --- |
| Age |  | 32 (24.0-41.5) | 27 (23.0-32.0) | 36 (32.0-40.0) | 33 (28-42) |
| Gender | Male | 46 (57.5%) | 8 (34.8%) | 0 (0.0%) | 36 (65.5%) |
|  | Female | 34 (42.5%) | 15 (65.2%) | 0 (0.0%) | 19 (34.5%) |
| Race | African | 79 (78.8%) | 23 (29.1%) | 1 (50.0%) | 55 (69.6%) |
|  | Indian | 1 (1.3%) | 0 (0.0%) | 1 (50.0%) | 0 (0.00%) |
| Relationship status | Single | 60 (75.0%) | 17 (73.9%) | 1 (50.0%) | 42 (76.4%) |
|  | Relationship | 17 (21.2%) | 6 (26.1%) | 0 (0.0%) | 11 (20.0%) |
|  | Married | 3 (3.8%) | 0 (0.0%) | 1 (50.0%) | 2 (3.6%) |
| Education completed | Primary school | 17 (21.3%) | 4 (17.4%) | 0 (0.0%) | 13 (23.6%) |
|  | High school | 61 (76.3%) | 19 (82.6%) | 1 (50.0%) | 41 (74.6%) |
|  | Diploma/degree | 2 (2.5%) | 0 (0.0%) | 1 (50.0%) | 1 (1.8%) |
| Employment | Employed | 24 (30.0%) | 9 (39.1%) | 1 (50.0%) | 14 (25.5%) |
|  | Self employed | 6 (7.5%) | 1 (4.4%) | 0 (0.0%) | 5 (9.1%) |
|  | In school | 5 (6.3%) | 1 (4.4%) | 0 (0.0%) | 4 (7.3%) |
|  | Unemployed | 45 (56.3%) | 15 (52.2%) | 1 (50.0%) | 32 (58.2%) |
| Mobile phone | No | 7 (8.8%) | 3 (13.0%) | 0 (0.0%) | 4 (7.3%) |
|  | Yes | 73 (91.3%) | 20 (87.0%) | 2 (100.0%) | 51 (92.7%) |
| Time since diagnosis | < 6 months | 25 (31.3%) | 22 (95.7%) | 0 (0.0%) | 3 (5.5%) |
|  | >= 6 months | 55 (68.8%) | 1 (4.4%) | 2 (100.0%) | 52 (94.6%) |

\*\*Unknown ART status: was not determined whether they were new initiates or had been on ART before
\*Re-engagement: re-engaging in care after a treatment interruption of at least 90 days

Exposure to the B-OK bottles led to a significant increase in treatment literacy among participants **(Figure 2)**. Prior to being introduced to the B-OK bottles, the median treatment literacy index score was 2 (IQR 0-3) out of 9. After being exposed to the B-OK bottles, treatment literacy was significantly higher (median score 7, IQR 5-7, p<0.001). The largest increases in treatment literacy were observed for questions about the effect of ART on improving one’s own health (16% to 95%) and on reducing transmission risk (14% to 94%). The only component of the literacy index that did not change significantly was knowledge about the need for immediate ART initiation following diagnosis (73% to 81%, p=0.13). The increase in treatment literacy following exposure to the B-OK bottles was significant both among participants newly initiating ART and those re-engaging in care; post-test scores in new initiates were higher compared to post-test scores for those re-engaging with care although the median scores were the same (7: IQR 7-7 among new initiates vs 7: IQR 5 -7 among re-engaging; p<0.001) (Figure 2). Prior to the B-OK bottle discussion, 70% of participants agreed that they “worried that HIV treatment did not completely eliminate the risk of transmission”, 14% disagreed, and 17% were not sure. After the bead bottle discussion, 65% of participants still agreed with this statement, while the percentage of those who disagreed increased to 33% and only 3% remained unsure. Exposure to the B-OK bottles also led to a significant increase in whether participants had ever heard of the terms “viral suppression” and “U=U”, from 20.0% to 98.8% and 8.75% to 98.8%, respectively **(Table A1)**

**Figure 2.**
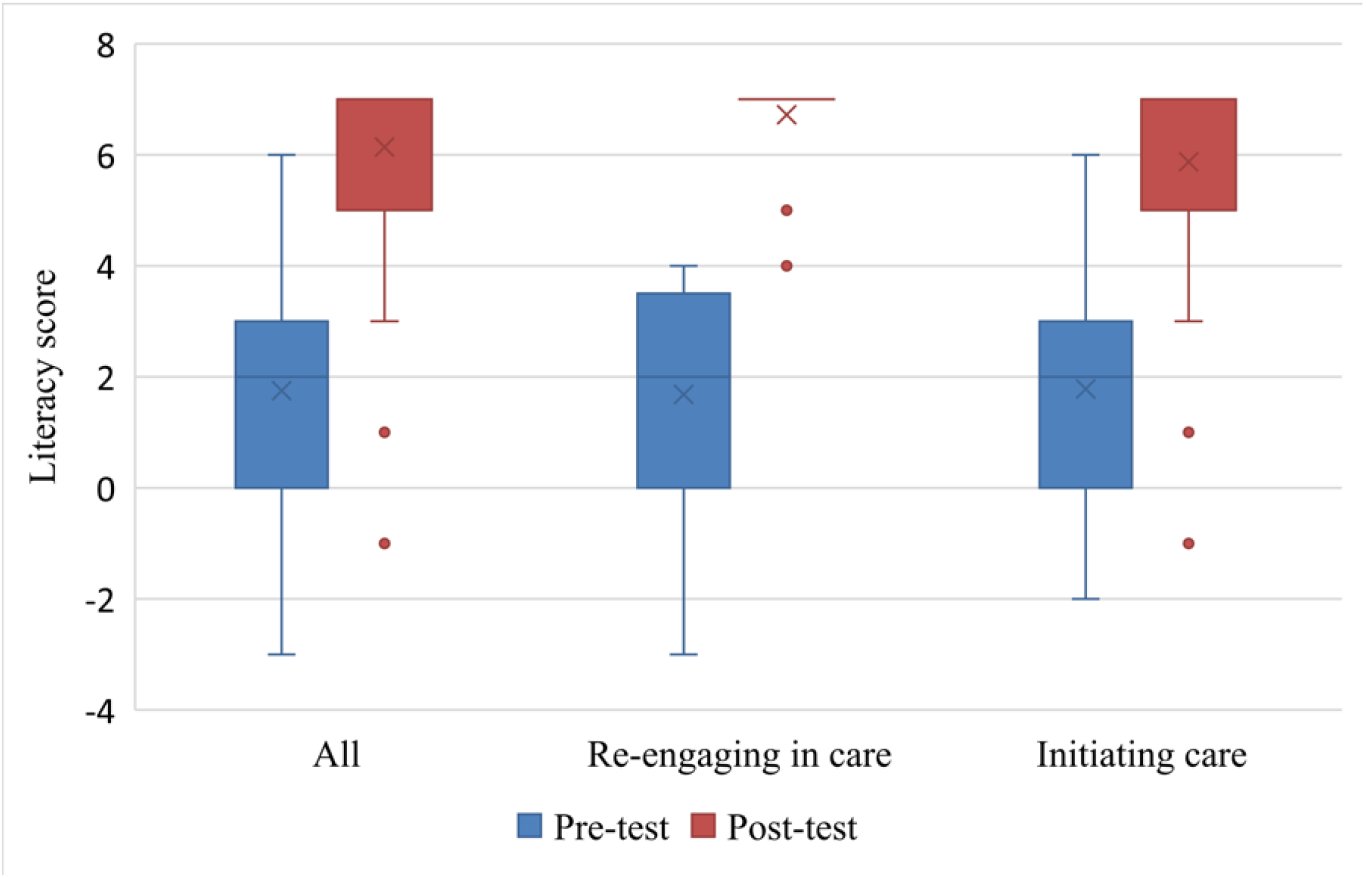
HIV treatment literacy scores before and after B-OK bead bottle discussion, by treatment status (re-engaging in care after a treatment interruption of at least 90 days or initiating treatment)

## Discussion

Use of visual aids in the form of coloured bead bottles to provide education about HIV and ART led to a large increase in treatment literacy among PLHIV at selected clinics in KwaZulu-Natal, South Africa. Data collected prior to use of the B-OK bottles confirmed findings from other studies (1,6) with participants having low treatment literacy, including limited awareness of the therapeutic and prevention benefits of ART. In contrast, following the short, 10-minute introduction to the B-OK bottles, care recipients reported much higher knowledge about the benefits of ART.

This study contributes to a growing body of research on strategies for increasing treatment literacy among PLHIV and the general population. While relatively few studies have been conducted in Sub-Saharan Africa (8,9), the existing evidence shows that provision of information about U=U in the form of flyers and interpersonal communication as well as community meetings were successful in increasing HIV testing uptake and reducing stigma (3). To our knowledge, this is one of the first studies to examine changes in treatment literacy through the use of simple visual aids that explain the benefits of ART.

Several studies have highlighted the difficulties in conveying the U=U message to PLHIV in a way that is easily understood (2,3,14). The B-OK bottles have the potential to remove some of these challenges. These bottles use visual aids, which have long been recognised as an effective way of communicating complex health information to, care recipients to explain concepts involving HIV in the body and the way that ART can reduce viral load and transmission risk. The bead bottles are also useful for explaining concepts involving probability. Enabling care recipients to engage with the bead bottles may have also facilitated their absorption of key health information relating to ART. Some advantages of the B-OK bottles include the low cost and ease with which they can be made and the short amount of time needed to use the bottles to explain HIV, ART and VLS. The bottles can also be used effectively by lay providers.

This study has several limitations. First, because there was no control group, changes in treatment literacy following the use of B-OK bead bottles may be confounded by other factors. Additionally, we measured the change in treatment literacy immediately after explaining the B-OK bead bottles, which limits our ability to assess whether care recipients’ increased treatment literacy was sustained over time and whether this impacted treatment uptake and adherence.

## Conclusions

This study shows that a simple visual aid that explains the effect of ART on HIV viral loads has the potential to improve awareness of the therapeutic and prevention benefits of ART among PLHIV. Future research should include rigorous, larger scale evaluations that determine the effect of the B-OK bottles and similar visual aids on not only treatment literacy, but also health behaviours and clinical outcomes among PLHIV. Implementation research that assesses strategies for introducing and adapting the B-OK bottles for large-scale use should also be a priority.

## Competing interests

The authors declare that they have no competing interests.

## Authors’ contributions

HT, SP, AB, CCM and SM conceptualized and designed the study. CG conducted the analysis. CG prepared the original draft. All authors reviewed and edited the draft.

## Author information

We would like to thank the all the participants and staff including data capturers and nurses at the facilities we conducted the research for their support and contribution to the success of the study.

## Acknowledgements

The authors gratefully acknowledge the contributions of the study participants and dedication of the staff at the study sites.

## Funding [Optional]

This analysis was funded by the Bill & Melinda Gates Foundation (INV-008318). The contents are the responsibility of the authors and do not necessarily reflect the views of the sponsor. The funders had no role in the study design, collection, analysis and interpretation of the data, in manuscript preparation or the decision to publish.

## Data Availability Statement [Mandatory]

The dataset analysed for the manuscript is available upon reasonable request.

## Supporting Information [Optional]

Supporting Information file 1:

**Table A1:**
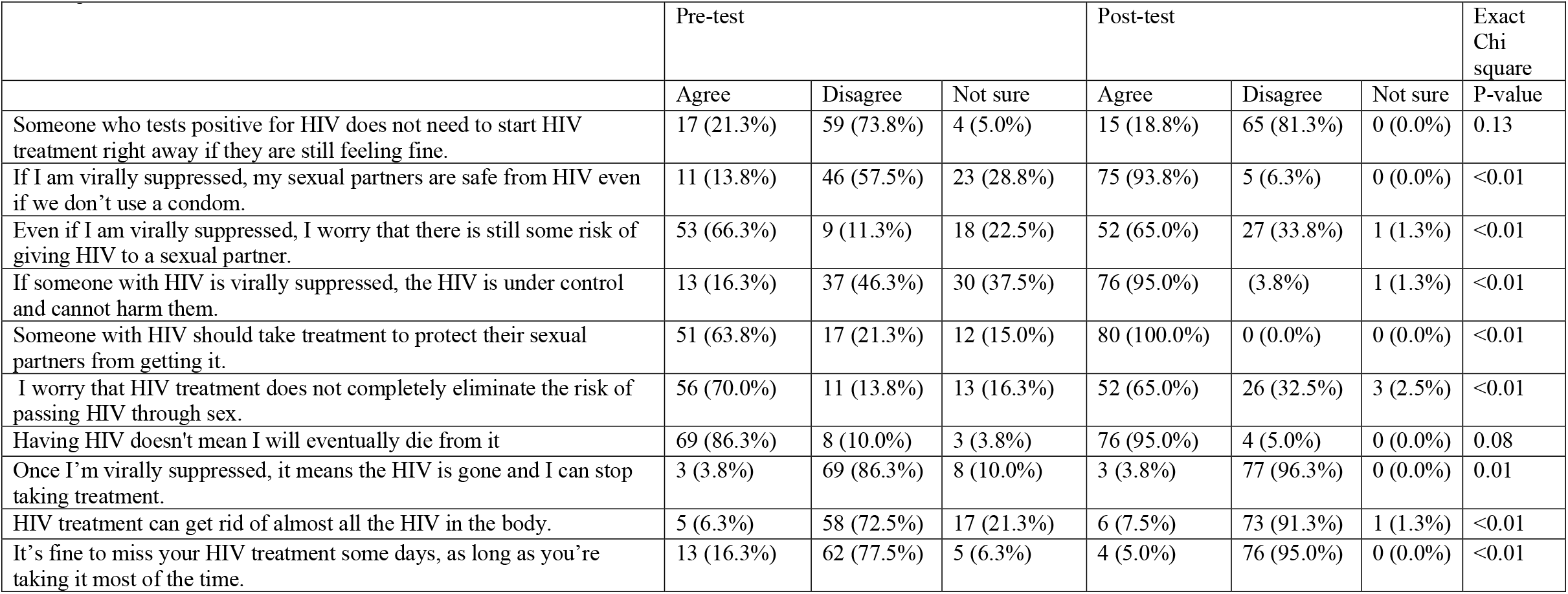
Comparison of HIV Treatment Knowledge Pre and Post Use of BOK Bead Bottles.

## Notes

### Competing Interest Statement

The authors have declared no competing interest.

### Clinical Trial

DOH-27-092022-8067

### Author Declarations

The ethics committees of University of the Witwatersrand (# M220707) and University of Pennsylvania (# 852087) gave ethical approval for this work.

### Summary of Updates

To add the appendix referenced in the article.

