## Appendix table 1 for "Increasing HIV treatment literacy among people living with HIV using a novel health communication aid: Evidence from KwaZulu Natal province, South Africa" (from another version of this preprint: version 1, 2023-06-20)

|  | Pre-test | | | Post-test | | | Exact Chi square |
| --- | --- | --- | --- | --- | --- | --- | --- |
|  | Agree | Disagree | Not sure | Agree | Disagree | Not sure | P-value |
| Someone who tests positive for HIV does not need to start HIV treatment right away if they are still feeling fine. | 17 (21.3%) | 59 (73.8%) | 4 (5.0%) | 15 (18.8%) | 65 (81.3%) | 0 (0.0%) | 0.13 |
| If I am virally suppressed, my sexual partners are safe from HIV even if we don’t use a condom. | 11 (13.8%) | 46 (57.5%) | 23 (28.8%) | 75 (93.8%) | 5 (6.3%) | 0 (0.0%) | <0.01 |
| Even if I am virally suppressed, I worry that there is still some risk of giving HIV to a sexual partner. | 53 (66.3%) | 9 (11.3%) | 18 (22.5%) | 52 (65.0%) | 27 (33.8%) | 1 (1.3%) | <0.01 |
| If someone with HIV is virally suppressed, the HIV is under control and cannot harm them. | 13 (16.3%) | 37 (46.3%) | 30 (37.5%) | 76 (95.0%) | (3.8%) | 1 (1.3%) | <0.01 |
| Someone with HIV should take treatment to protect their sexual partners from getting it. | 51 (63.8%) | 17 (21.3%) | 12 (15.0%) | 80 (100.0%) | 0 (0.0%) | 0 (0.0%) | <0.01 |
| I worry that HIV treatment does not completely eliminate the risk of passing HIV through sex. | 56 (70.0%) | 11 (13.8%) | 13 (16.3%) | 52 (65.0%) | 26 (32.5%) | 3 (2.5%) | <0.01 |
| Having HIV doesn't mean I will eventually die from it | 69 (86.3%) | 8 (10.0%) | 3 (3.8%) | 76 (95.0%) | 4 (5.0%) | 0 (0.0%) | 0.08 |
| Once I’m virally suppressed, it means the HIV is gone and I can stop taking treatment. | 3 (3.8%) | 69 (86.3%) | 8 (10.0%) | 3 (3.8%) | 77 (96.3%) | 0 (0.0%) | 0.01 |
| HIV treatment can get rid of almost all the HIV in the body. | 5 (6.3%) | 58 (72.5%) | 17 (21.3%) | 6 (7.5%) | 73 (91.3%) | 1 (1.3%) | <0.01 |
| It’s fine to miss your HIV treatment some days, as long as you’re taking it most of the time. | 13 (16.3%) | 62 (77.5%) | 5 (6.3%) | 4 (5.0%) | 76 (95.0%) | 0 (0.0%) | <0.01 |

**Table A1: Comparison of HIV Treatment Knowledge Pre and Post Use of BOK Bead Bottles**
